# TSABL: Trait Specific Annotation Based Locus Predictor

**DOI:** 10.1101/2021.09.27.21264198

**Authors:** Kimberly Lorenz, Christopher S. Thom, Sanjana Adurty, Benjamin F. Voight

## Abstract

The majority of GWAS loci fall in the non-coding genome, making causal variants difficult to identify and study. We hypothesized that the regulatory features underlying causal variants are biologically specific, identifiable from data, and that the regulatory architecture that influences one trait is distinct compared to biologically unrelated traits. To better characterize and identify these variants, we used publicly available GWAS loci and genomic annotations to build 17 Trait Specific Annotation Based Locus (TSABL) predictors to identify differences between GWAS loci associated with different phenotypic trait groups. We used a penalized binomial logistic regression model to select trait relevant annotations and tested all models on a holdout set of loci not used for training in any trait. We were able to successfully build models for autoimmune, electrocardiogram, lipid, platelet, red blood cell, and white blood cell trait groups. We used these models both to prioritize variants in existing loci and to identify new genomic regions of interest. We found that TSABL models identified biologically relevant regulatory features, and anticipate their future use to enhance the design and interpretation of genetic studies.

## Introduction

Genome-wide association studies (GWAS) are a widely used method to identify genomic regions associated in a population with a phenotype of interest, and have successfully detected thousands of loci across the human genome, each of which contains at least one causal variant^1–16^. Unfortunately, identifying the causal variant(s) and underlying mechanism at associated loci is far more challenging, and for many genomic regions there exist more potential causal variants in credible sets than can be experimentally tested^17–19^. While trans-ancestry fine-mapping can help to narrow the set of putative variants^20^, this approach is limited to loci which share genetic architecture and genetic variation across populations, and still may not resolve causal variant(s) within a locus. Therefore, an orthogonal method for prioritizing variants in GWAS loci would be beneficial. Additionally, as more well-powered GWAS reveal more loci, a straightforward method to prioritize the likelihood of variants contributing to a specific disease will aid in locus discovery.

While pleiotropy is pervasive across the human genome^21^, the contribution of individual variants depends on the biological relatedness of the traits under consideration. For example, the loci, causal genes, and variants associated with circulating plasma lipid levels might reasonably be expected to differ from those that modify electrocardiogram (ECG) traits, as the biological basis for these traits involve different tissues (liver vs. heart), different physiologies (lipid metabolism and trafficking vs. cardiac electric conductance), and different genes (e.g., *SORT1*^22^ vs. *NOS1AP*^23^). Presumably, then, with sufficient association data and a diverse and dense collection of functional annotations across the genome, it might be possible to train a model to predict loci – and perhaps even variants – that relate to *specific* sets of traits relative to all others. Ideally, such a predictor would learn and select biologically relevant annotations from a set of all possible genomics features, thereby also providing informative hypotheses for why particular loci or variants are predicted to be “trait-relevant”. Current methods to predict if a variant is likely functional for *any* trait (such as CADD^24^, GWAVA^25^, and DeepSEA^26^) provide a score based on likelihood of function across multiple (unrelated) phenotypes and diseases.

Previously, we built platelet and red blood cell models using those GWAS as positives and random matched genomic regions as controls^27^. While this was successful, we found the models to not be trait selective. Here, we instead sought to develop a trait specific annotation based locus (TSABL) predictor which would discriminate loci associated with a specific trait (or collection of related traits) from all other complex trait associated loci identified by GWAS. We collected association results from 42 GWAS-scanned phenotypes^1–16^, coarsely grouping them into 17 related trait groups to test and validate prediction models (**Supplementary Table 1**). We then applied a penalized binomial logistic regression model (LASSO) to each trait group using publicly available genome wide features such as DNase hypersensitivity and histone or transcription factor ChIP-seq (**Supplementary Table 2**), along with gene expression, phastCon score, and size of locus. The six successful TSABL models were able to separate regions associated with their trait group from other GWAS loci and identified sensible biological features for the phenotypes in focus. We then applied these models to fine mapping and locus discovery paradigms, both of which may lead to translationally relevant trait-specific biological mechanisms.

## Results

To develop our approach, we needed to define the set of traits, identify positive and negative loci for those traits, and obtain a collection of genomic features to discriminate between these labels. For traits to study, we focused on 42 phenotypes obtained from curated GWAS and the NHGRI-EBI GWAS catalog that we collapsed into 17 related trait groupings (**Methods, Supplementary Table 1**). Using all GWAS lead variants identified, we first clumped variants into loci by linkage disequilibrium (using European LD R^2^ > 0.7). Next, for each trait group we defined positive loci as those which contained at least one variant with an associated *P*-value < 5 × 10^−8^ with at least one of the phenotypes in the trait group. Negative loci for each trait group were defined as those without any variant even modestly associated (*P*-value > 0.05) with one or more of the phenotypes in the trait group. We utilized genome-wide annotations from multiple public resources, including nearest gene tissue-specific expression and epigenomic features, tagging these features across positive and negative examples if the features overlapped with any variant in the locus.

We then generated predictive models for each of the 17 trait groups using a penalized binomial logistic regression model (LASSO)^28^, using cross-validation for training and estimating final prediction accuracy on a holdout set not used in training any model (**Methods, Supplementary Figure 1**). We defined a model as informative if the area under the curve (AUC) was ≥ 0.7 and the difference between training and holdout AUCs was ≤ 0.02 (implying only a small level of overfitting). Based on these criteria, models trained for autoimmune, electrocardiogram (ECG), lipid, platelet, red blood cell (RBC), and white blood cell (WBC) traits passed, and were the focus of further investigation (**Figure 1a, Table 1**).

**Figure 1:**
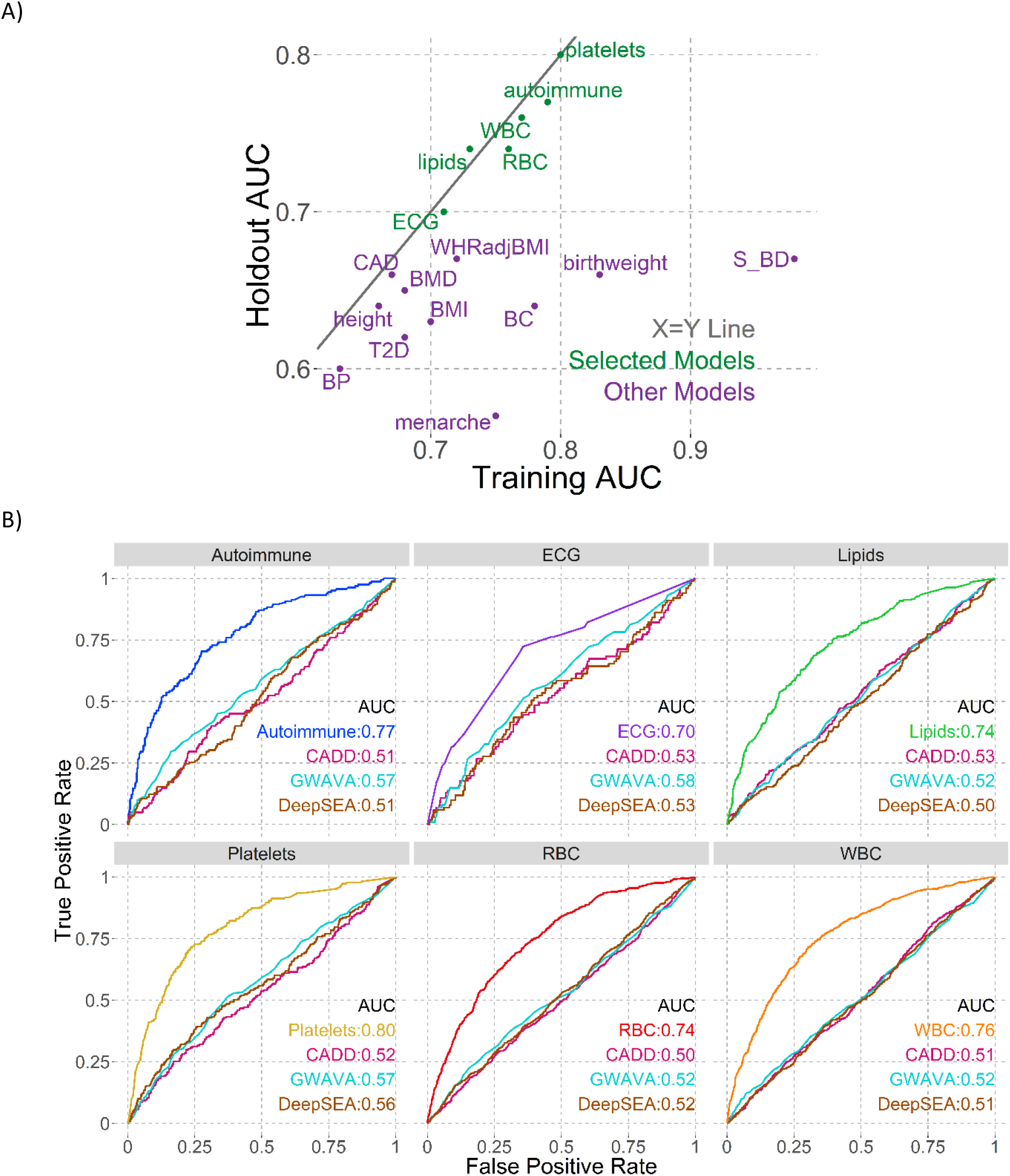
Model AUCs. A) Training vs Holdout AUCs. Dotted grey line shows equality. Selected models (holdout AUC ≥ 0.7) shown in green, unselected in purple. B) Individual holdout AUC plots for selected models, compared to CADD (pink), GWAVA (cyan), DeepSEA (brown).

**Table 1:** All Traits Results Summary.

| Trait | Positive SNPs | Total Positive Loci | Positive Training Loci | Positive Holdout Loci | Training AUC | Model Holdout AUC | CADD Holdout AUC | GWAVA Holdout AUC | DeepSEA Holdout AUC | Number Selected Features |
| --- | --- | --- | --- | --- | --- | --- | --- | --- | --- | --- |
| platelets | 955 | 699 | 464 | 231 | 0.80 | 0.80 | 0.52 | 0.57 | 0.56 | 17 |
| autoimmune | 724 | 484 | 318 | 162 | 0.79 | 0.77 | 0.51 | 0.57 | 0.51 | 37 |
| WBC | 2222 | 1762 | 1091 | 544 | 0.77 | 0.76 | 0.51 | 0.52 | 0.51 | 26 |
| lipids | 1439 | 810 | 526 | 275 | 0.73 | 0.74 | 0.53 | 0.52 | 0.50 | 15 |
| RBC | 2430 | 1810 | 1132 | 564 | 0.76 | 0.74 | 0.50 | 0.52 | 0.52 | 31 |
| ECG | 444 | 303 | 202 | 101 | 0.71 | 0.70 | 0.53 | 0.58 | 0.53 | 4 |
| S_BD | 624 | 422 | 273 | 144 | 0.98 | 0.67 | 0.48 | 0.49 | 0.45 | 519 |
| WHRadjBMI | 506 | 419 | 275 | 141 | 0.72 | 0.67 | 0.54 | 0.56 | 0.56 | 5 |
| birthweight | 346 | 294 | 195 | 97 | 0.83 | 0.66 | 0.48 | 0.59 | 0.53 | 75 |
| CAD | 740 | 535 | 346 | 183 | 0.67 | 0.66 | 0.52 | 0.54 | 0.53 | 7 |
| BMD | 3046 | 2140 | 1394 | 697 | 0.68 | 0.65 | 0.53 | 0.52 | 0.52 | 42 |
| BC | 419 | 328 | 218 | 109 | 0.78 | 0.64 | 0.53 | 0.47 | 0.54 | 53 |
| height | 3715 | 3392 | 1699 | 839 | 0.66 | 0.64 | 0.51 | 0.50 | 0.52 | 24 |
| BMI | 2220 | 1676 | 1032 | 516 | 0.70 | 0.63 | 0.50 | 0.51 | 0.51 | 53 |
| T2D | 849 | 587 | 388 | 194 | 0.68 | 0.62 | 0.51 | 0.53 | 0.54 | 18 |
| BP | 2595 | 1842 | 1215 | 616 | 0.63 | 0.60 | 0.50 | 0.51 | 0.51 | 10 |
| menarche | 817 | 593 | 393 | 197 | 0.75 | 0.57 | 0.52 | 0.44 | 0.49 | 59 |

To assess why these 6 traits produced successful models while the other 11 failed, we considered whether trait models with more positive loci or more features were more predictive (**Supplementary Figure 2**). We did not find an obvious relationship between either number of loci used to train models or number of annotations selected and holdout set AUC, suggesting that our inability to predict these traits is not based on limitations in availability of positive/negative examples or the total number of available features. The implication may be that the catalog of existing features available for modelling insufficiently captures differences between these other 11 traits and the spectrum of complex traits more generally.

We considered two metrics of model specificity. First, trait specific models should be more informative for prediction relative to functional variant prediction methods which were designed to be trait agnostic. For the trait groups we assembled, the holdout set AUCs for CADD, GWAVA, and DeepSEA were all close to 0.5, indicating that none were informative for discriminating amongst trait-associated GWAS loci (**Figure 1B, Table 1**). Second, trait specific predictors should perform the best on the phenotype collections from which they were trained. Looking at comparative model performance, we observed that all six selected models were better at predicting their own holdout set compared to others (**Figure 2, left panels** and **Supplementary Figure 3**). These results indicate that these six prediction models can discriminate between traits using GWAS loci alone, and that the power to make predictions is not due to intrinsic functionality of GWAS loci in general that would be captured by trait agnostic methods.

**Figure 2:**
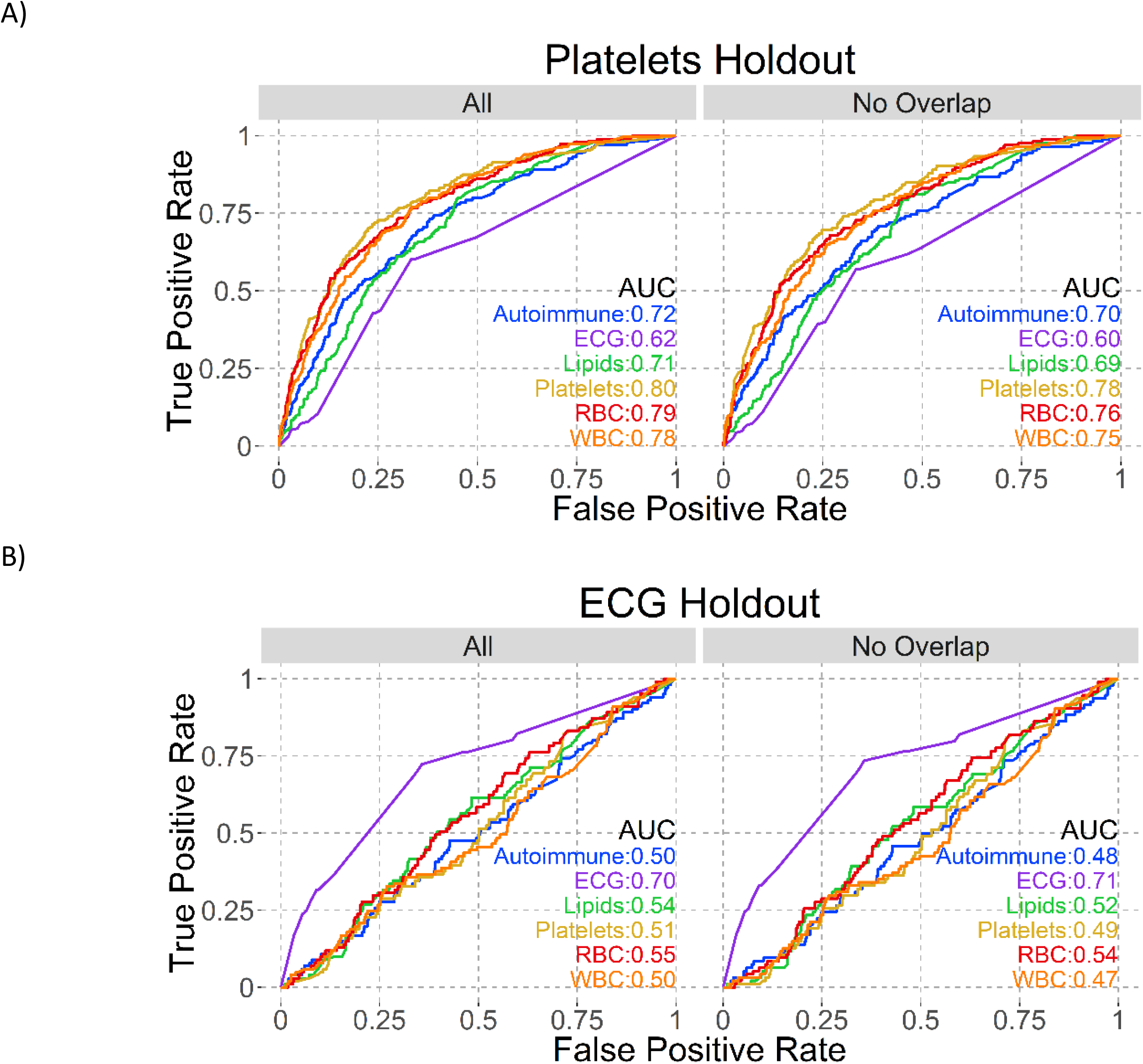
Model AUCs on same holdout sets. Each panel shows the named holdout set: A) Platelets and B) ECG. AUCs for all six selected models are shown on all plots, with the left (All) plot having all loci in that holdout and the right (No Overlap) having only those loci not positive in multiple models.

Given the pervasive nature of pleiotropy, we would expect that predictors for traits that are related to one another – but perhaps not so closely as to be grouped together – should be partially cross-predictive. We observed this to be true for the RBC model, which could be used to predict the platelet holdout set with an AUC of 0.79 (**Figure 2A, left panel**). To rule out that this similarity was driven by loci that are positive for both traits, we removed all loci from the holdout set that were positive in both the “in-focus” trait (here, platelets) and any other model, then reassessed the holdout AUC for each model. Even after removing overlapping loci, the RBC model still predicted the platelet holdout set well, indicating the prediction accuracy is not driven by overlapping loci (RBC AUC of 0.76 on platelets holdout set; **Figure 2A, right panel**). In contrast, the ECG model was the only one that predicted the ECG holdout set reasonably, while the autoimmune, lipids, platelets, RBC and WBC models all had AUC < 0.55 on the ECG holdout (**Figure 2B**).

Looking at the annotations selected as important for our six successful models, we found them to be largely in line with known trait biology (**Figure 3, Supplementary Figure 4, Supplementary Table 3)**. All three blood traits (platelets, RBC, and WBC) selected mainly whole blood annotations that positively rank active regions (active histone marks, gene expression, and transcription factor (TF) binding) and negatively rank repressive histone marks (H3K9me3) (**Figure 3A, Supplementary Figure 4B and 4C**). By contrast, the ECG model selected only heart tissue annotations (**Figure 3B**), and the lipids model chose predominantly liver tissue features (**Figure 3C**). We also identified features that distinguish these traits from each other. For example, only the models for the immune traits autoimmune and WBC selected spleen gene expression.

**Figure 3:**
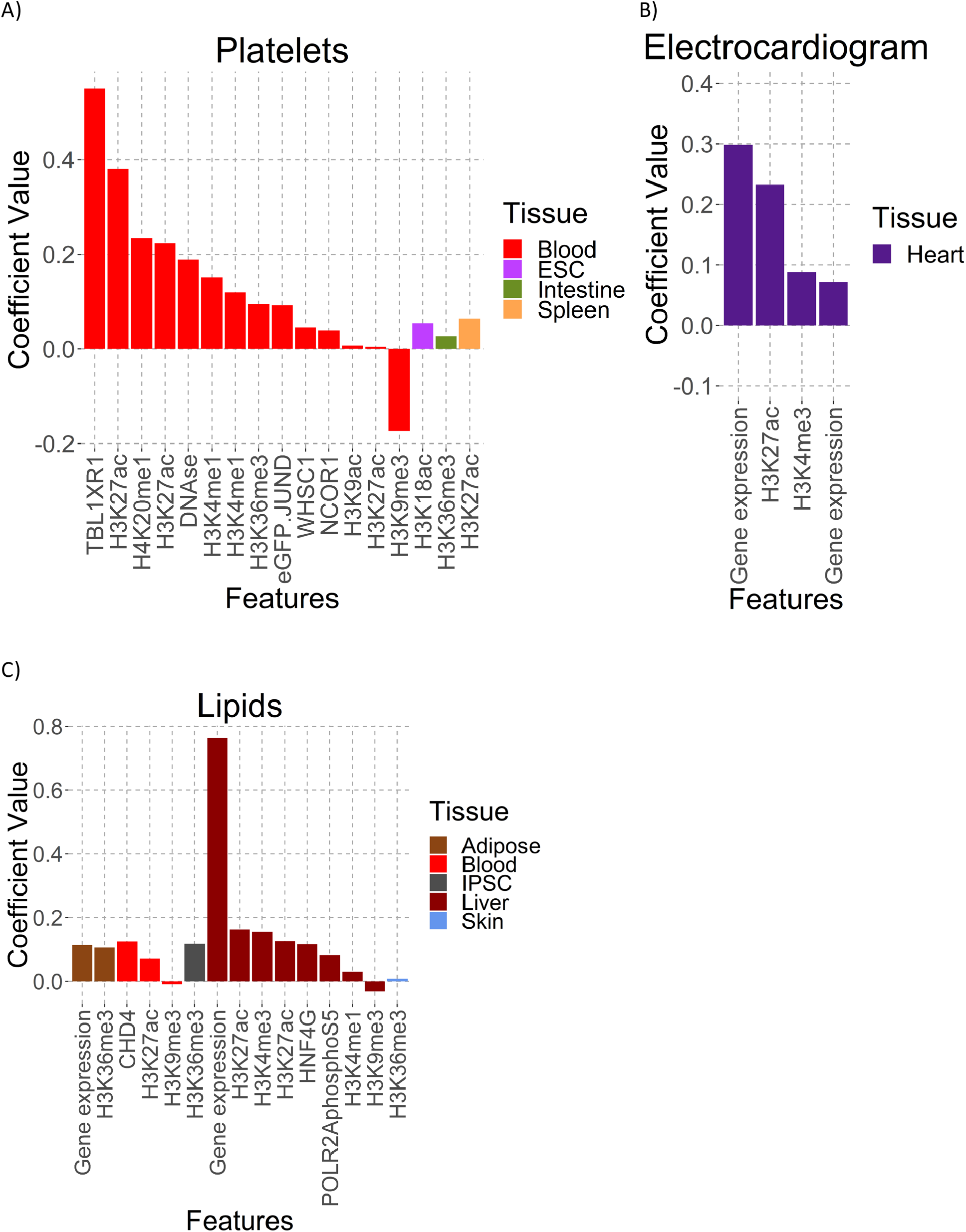
Selected Model Feature Coefficients. A) Platelets, B) ECG, and C) Lipids model coefficients, sorted by tissue type and value.

We next used the framework of stratified LDScore regression^29^ to evaluate if the scores emitted by our models could be used as an annotation to explain partitioned heritability. With our six added annotations derived from our six selected models, we tested 103 annotations in total and so considered enrichments with *P*-value < 4.8 × 10^−4^ significant by Bonferroni correction. We found enrichment in GWAS for all successful models with summary statistics available (autoimmune, lipids, platelets, RBC, WBC) (**Supplementary Table 4**). To test whether this enrichment was driven by the lead GWAS SNPs that the models were based on, we also removed these lead GWAS SNPs and their LD proxies (EUR R^2^ LD > 0.7) from the dataset and reassessed enrichment. We found that the lipids, platelets, and WBC models were significant in all individual phenotype association data, and the RBC model was significant for 3 of 4 phenotype GWAS (**Supplementary Table 5**). Perplexingly, the autoimmune model was not significant for any phenotype GWAS after lead SNPs were removed, but the related trait models for WBC and platelets were significant for all 3 autoimmune phenotype GWAS. We expect that the autoimmune trait group failed this test due to a lack of power, as it had the fewest positive loci of the tested successful models. The enrichment we see in LDScore partitioned heritability demonstrates that our trait- and cell type-specific modeling approach can define target loci for related disease phenotypes, facilitating downstream validation and biological/mechanistic understanding.

### Application - Locus Discovery

We next applied our six predictive models to all SNPs identified in phase III of the 1000 Genomes project. From ∼78 million scored SNPs, we predicted between 7,366 and 485,424 trait associated SNPs per model that exceeded a FDR < 0.01 threshold (**Supplementary Table 6**). To find genes associated with these SNPs, we first identified the 436 and 2,137 genes overlapping these variants, of which 153 to 879 were outside of identified GWAS regions (defined as 1MB window surrounding lead SNPs; **Supplementary Table 6**). For example, the lipids model identified 18,847 SNPs, with 458 overlapping genes, of which 167 were outside known GWAS regions. While we found many SNPs near established GWAS variants, we were interested here in the regions not previously identified by GWAS analyses, and our models highlight several points of potentially interesting biology. For the lipids model, we identified a cluster of 10 SNPs overlapping *DHCR24*, a known cholesterol biosynthesis gene^30^, and a cluster of 63 SNPs overlapping *PLIN2*, a gene involved in lipid globule storage^31^, both of which are reasonable biological candidates for lipid related phenotypes. For the platelets model, we identified a cluster of 8 SNPs overlapping *JARID2*, a gene known to be involved in hematopoiesis^32^, and 7 SNPs overlapping *PXN*, which is involved in immunity response^33^, both of which may plausibly impact platelet phenotypes. Additionally, we found that FDR < 0.01 variants of the platelets, RBC, and WBC models were enriched for being within 25kb of a newly published set of blood traits GWAS^34^ (**Supplementary Figure 5**).

To assess whether the trait associated SNPs mapped onto established biological knowledge, we used GREAT^35^ analysis to examine both the human gene ontology (GO) biological process and mouse single knockout (KO) phenotypes associated with these SNPs. We used both the full set of trait associated SNPs and also removed all variants within 500 KB of identified GWAS regions (non-GWAS variants) (**Supplementary Table 7**). For the full set of trait associated SNPs, all models returned mouse KO phenotypes consistent with our expectations. For example, mouse KOs for genes overlapping our autoimmune associated SNPs are enriched for phenotypes such as “abnormal T cell morphology” and “decreased lymphocyte cell number.” GO biological processes were similarly consistent with expectations for autoimmune, ECG, lipids, and WBC trait models, while the platelets and RBC traits returned mostly generic results (for example, “RNA processing”). The non-GWAS variants analyses were similar, except that the lipids and RBC models did not return any results for either the mouse KO or GO biological processes analyses.

### Application – Fine Mapping

Functional cellular follow-up studies require one to prioritize GWAS loci, genes and variants within them that are likely causal. To assess the usefulness of our model scores for individual SNPs within GWAS loci, we identified SNPs in credible sets that exceed FDR thresholds of 0.10, 0.25, or 0.50 of the relevant model for phenotypes where appropriate summary statistics were available (autoimmune, platelets, RBC and WBC, **Table 2, Supplementary Tables 8 and 9**). At FDR 0.10, our models reduced the size of credible SNPs of interest in 27% of multi-SNP credible sets; at FDR 0.25, 52% of credible sets see a reduction in SNP number. For many of these loci the reduction is substantial. For example, lead rs10486483 from Crohn’s Disease had a credible set of 85 variants that was reduced to 3 after applying our FDR 0.10 threshold; or lead rs2382817 in Inflammatory Bowel Disorder, where a credible set of 68 was reduced to 5 variants at the same threshold. While a set threshold is useful for summarizing, it is not necessary in practice, and variants in a credible set can simply be ordered by their TSABL score for assessment, facilitating immediate integration into functional follow up study pipelines.

**Table 2:**
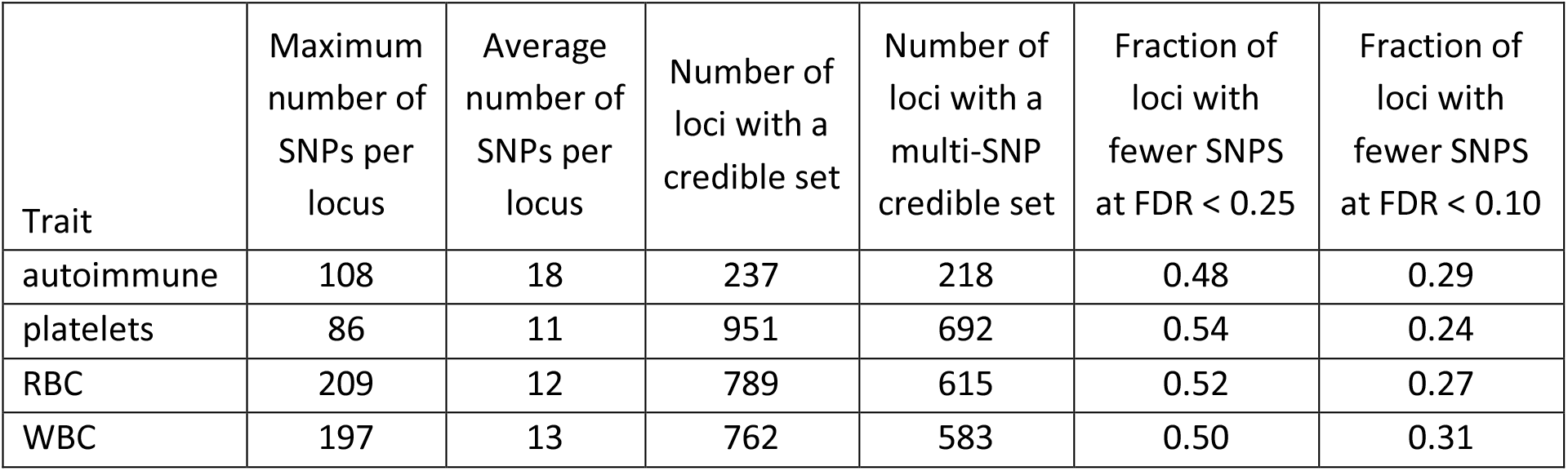
Credible set improvements.

## Discussion

With current pitfalls in fine mapping and locus discovery, our approach can distinguish amongst complex trait-related groups applied to 17 sets of traits. Our modeling process, built using publicly available GWAS and annotation data, selects and outputs features and variants relevant to underlying disease biology. These models predict differences between traits that currently available SNP ranking schemes do not, and the overlaps we see among models are consistent with known biological links between the traits assessed. Our modelling strategy makes no assumptions about number of causal variants in a GWAS locus, is expandable to include new genome wide features as they are developed, and can be applied to locus discovery problems or to prioritization after fine-mapping for functional validation efforts.

This approach is limited by both the GWAS data and genome wide features available. Surprisingly, we were unable to adequately model some complex traits (**Table 1**). However, we expect that additional genomic features will improve some models; for example, in the dataset used for this analysis there are 642 annotations from blood tissue but only 31 from bone (**Supplementary Table 2**) and it seems likely that part of the reason the height model was not well specified is that it lacked key genomic annotations relevant to bone growth. Additionally, the rapidly expanding repertoire of single cell data sets may better parse complex tissue types (e.g., pancreas) and capture data relevant to rare cell types that are underrepresented in current datasets. Future studies will be designed to incorporate these data types, which are only currently emerging.

We selected a modelling strategy to allow for improved biological interpretability, but this presumably comes at a cost of accuracy. We intentionally selected a variety of traits with very different underlying physiology, and we found that the traits well described by our models are linked largely to single cell types, namely blood, liver, or heart. We do not successfully model traits such as Body Mass Index (BMI) or Type 2 Diabetes (T2D), both of which involve many tissues and have disease subtypes that are not well captured in current GWAS analyses^36–38^. We expect that analyzing these phenotypes as a single cohort may serve to obfuscate rather than clarify, and anticipate that incorporating more granularity in phenotypes (type of T2D or severity of BMI, for example) will improve model performance.

Pleiotropy presents another challenge to our modeling approach, but is inherently captured in any genomic data set. It is plausible that more sophisticated approaches with this in mind might be anticipated to improve performance (e.g., multi-label classification). Moreover, approaches that captured uncertainty around labelled examples (e.g., “noisy” labels) might also be helpful to address these challenges as well as expand the use of both positive and negative examples. Additional processing of genome wide features, both to account for correlation and to reduce the feature space prior to model building (e.g., stability selection) may also improve results.

A logical point of comparison for TSABL is to another strategy used for fine mapping or locus discovery. The most similar is the fGWAS method, which is designed to jointly consider GWAS and genomic annotations^39^. The main distinction between these models is that fGWAS considers trait loci compared to all regions of the genome (regardless of their association with a complex trait) while TSABL specifically uses established GWAS loci as the negative set. In practice, what this means is that TSABL models found differences between traits, rather than annotations that may be held in common between all GWAS loci. While both approaches are useful, identifying trait specific annotations will allow for the translation of GWAS loci into functional biological hypotheses, facilitating disease treatment and variant-based drug discovery.

## Methods

### General data sources and analysis

All chromosome:position coordinates given in this paper refer to genome build hg19. We downloaded refseq gene exon coordinates for build hg19 from ENSEMBL^40^ at http://grch37.ensembl.org/biomart/ on 10/24/17, and used the extreme exon boundaries as gene boundaries. We assessed annotations for rsIDs of interest using ANNOVAR version 2016Feb01^41^ and regulomeDB^42^.

### GWAS trait grouping

Full listing of GWAS phenotypes included in each of the 17 trait groups are found in **Supplementary Table 1**. We grouped GWAS together based on known biological links. The traits that include multiple GWAS analysis are autoimmune, blood pressure, electrocardiogram (ECG), lipids, platelets, red blood cell (RBC), type 2 diabetes (T2D), and white blood cell (WBC), For autoimmune, we used the five most correlated traits (psoriasis, Crohn’s disease, ulcerative colitis, Behcet’s disease and ankylosing spondylitis) in an analysis of autoimmune disease correlation^43^ plus inflammatory bowel disorder due to its similarity to ulcerative colitis and Crohn’s disease^10^. For T2D, we also included fasting glucose and fasting insulin results from the NHGRI-EBI catalog, under the reasoning that either or both are used in the clinical diagnosis of T2D. For the remaining trait groups, all GWAS phenotypes included were assessed in the same paper and are highly correlated.

### GWAS loci identification & processing

We downloaded the NHGRI-EBI GWAS catalog v1.0 2019-06-20 from https://www.ebi.ac.uk/gwas/docs/file-downloads on 06/21/19 and processed GWAS results by removing the following associations:

1. with no listed *P*-value or a *P*-value > 5e-8
2. with no rsID listed or multiple rsIDs listed
3. listed as on the X chromosome, no chromosome, or multiple chromosomes
4. with descriptions indicating they were interaction effects with another locus
5. located in the HLA region (chr6:29670261-33104175; hg19 coordinates)

Remaining associations were grouped by rsIDs, resulting in 53,640 variants.

We also included lead variants from GWAS for specific phenotypes associated with our 17 trait groups, regardless of their inclusion in the GWAS catalog (autoimmune^3,10^, breast cancer^14^, birthweight^6^, bone mineral density^15^, body mass index^8^, blood pressure^11^, coronary artery disease^4^, electrocardiogram^5^, height^8^, lipids^12^, age at menarche^9^, platelets^1^, red blood cells^1^, schizophrenia/bipolar disorder^2^, type 2 diabetes^13^, white blood cells^1^, and waist hip ratio adjusted for BMI^16^; **Supplementary Table 1**). Literature curated GWAS variants were processed in the same way as catalog variants so that our input was a list of variant rsIDs.

For all variants, we identified hg19 positions using Ensembl biomart^40^, and removed variants if they could not be assigned a position. We identified loci of interest by collecting all variants with R^2^ ≥ 0.7 to a GWAS variant in European 1000 genomes phase III^44^ data using PLINK 1.90Beta4.5^45^. Variants were considered part of a locus if they had R^2^ ≥ 0.7 with any other variant in the locus. Final variant list includes 715,404 variants assigned to 32,713 loci and can be found in **Supplementary Table 10**.

### Feature identification and processing

Gappedpeak files for histone ChIP-seq experiments and the “hotspot.fdr0.01.broad” files for DNase assays were downloaded on 02/10/16 from the consolidated epigenomes section of the Roadmap Epigenomics Project^46^ portal. Uniform DNase files were downloaded 03/28/16 from http://hgdownload.cse.ucsc.edu/goldenPath/hg19/encodeDCC/wgEncodeAwgDnaseUniform/. Homo sapiens FAIRE, transcription profiling by array, and ChIP-seq data were downloaded 01.06.16 from the ENCODE^47^ project portal. We included data as provided for experiment accession numbers with a single file, and used the intersection if multiple files were provided. For transcription profiling by array data, we selected data labeled as “filtered transcribed fragments” if available. For ChIP-seq files, if the experiment accession had a file labeled “optimal idr threshold” or “replicated peaks” we used that file. For histone data, if both broadpeak and narrowpeak files were available, we created gappedpeak files. If only one peak type was available, we included that file. For all ChIP-seq targets that were not histones, narrowpeak files were used if available and broadpeak files otherwise. The complete list of 2305 ENCODE and Roadmap Epigenomics features used in this paper is found in **Supplementary Table 2**.

Feature overlaps with GWAS loci were identified using bedtools2v2.25.0 intersect^48^. For a given locus, a feature was coded as 1 if any variant in the locus overlapped the feature, and a 0 if not. For indel variants, overlap was counted only with the start position of the variant.

To build the features for tissue-specific gene expression, we utilized previously calculated t-statistics for specific expression^49^. The nearest gene to each locus was determined as the gene containing a corresponding tissue-specific expression t-statistic with a start position nearest to the locus center. The locus was assigned a 1 in a tissue if the nearest gene had a t-statistic ranking in the top ten percent for that tissue and a 0 otherwise, creating 53 tissue-specific nearest gene expression features.

We downloaded 100way phastCon^50^ scores for the human genome on 04/26/16 from http://hgdownload.soe.ucsc.edu/goldenPath/hg19/phastCons100way/ and used the bigWigAverageOverBed^51^ tool from the UCSC toolkit to extract scores for single SNPs. We use the highest variant 100way phastCon score as the score for the locus.

Finally, we counted the number of variants in a locus and included normalized variant number as a possible feature.

Tables of features used for modelling are in **Supplementary Table 11**.

### Benchmarking scores

We downloaded CADD^24^ scores for 1000 genomes phase 3 variants from http://cadd.gs.washington.edu/download on 02/25/16. We downloaded GWAVA^25^ scores on 02/25/2016 from ftp://ftp.sanger.ac.uk/pub/resources/software/gwava/v1.0/annotated/GWAVA_db_csv.tgz and used tss_score for all analyses. For both CADD and GWAVA, a higher score indicates a greater probability of functionality, so if the downloaded database had multiple scores for a position, we used the highest score provided. If they did not have a score for a position, it was set to 0. We use the highest variant score as the locus score for both CADD and GWAVA.

We used the standalone deepSEA-0.94^26^ program to get deepSEA scores for our variants. Unlike the previous scores, a smaller e value corresponds to a greater probability of functionality, so we use the lowest variant deepSEA score as the score for the locus. For comparison purposes, we subtract this value from 1 so that it is on the same scale as the other metrics.

### Building models

For model training, we considered a locus as being positive for a trait if at least one variant in the locus was associated with the trait and negative otherwise. We separated all loci into 2/3 training and 1/3 holdout. This separation was done semi-randomly, with the following conditions enforced: 1) for each trait, maintained a 2/3 train, 1/3 holdout ratio for positive loci, and 2) maintained native distribution of variant count in loci (for example if there were 60 loci containing 10 variants each, 40 were assigned to training and 20 to holdout). Furthermore, we removed any locus with > 1000 variants (1 locus removed). In this way we created a separate set of holdout loci that were not used to build any of the trait models and so could be used to compare model predictions across traits.

For each trait, we processed the training and holdout sets to identify positive and negative loci. First, we identified positive loci where at least one variant had *P*-value < 5 × 10^−8^ with a phenotype in the trait group. From the remaining loci, we removed those with *P*-value < 0.05 in the associated GWAS (if summary statistics were available – no removals for ECG or psoriasis GWAS were possible) to create a negatives pool. Finally, we used only positive loci where we could match 6 (5 for height) negative loci with similar locus variant counts: within 10 of a locus with variant count < 100 or within 50 of a locus with variant count >= 100. This ensured that our positive and negative locus pools were roughly matched for locus size. Using these processed positive and negative locus sets for each trait, we built models using the glmnet^28^ package in R, using binomial logistic regression with LASSO regularization and maximizing the AUC. The feature pool included 2305 ENCODE and Epigenome Roadmap annotations, 53 tissue-specific nearest gene expression annotations, the 100way Phastcon score, and normalized variant number. We used 10-fold cross validation to select lambda.s1e, performed 15 trials of model building, and selected the median training AUC model as the trait model for analysis (**Supplementary Figure 1)**.

### Model analysis

General model performance was assessed using the ROCR package^52^ in R on the holdout data & compared to the GWAVA, CADD, and DeepSEA predictors. We considered a model successful if the holdout AUC was > 0.7 and the difference between training and testing AUC was < 0.02; this includes models describing autoimmune, ECG, lipid, platelet, RBC, and WBC traits. Trait specific model performance was assessed by comparing the six selected model predictions on each holdout set. FDR thresholds used in further analysis were calculated from holdout datasets only.

Due to the nature of GWAS, there are loci tagged as positive across multiple traits. It is possible that these loci were driving the correlation between models for different traits. To test this, we removed all loci that are positive in the holdout set for the trait of interest and any other well modeled trait, creating a non-overlapping holdout set. We then compared ROC plots of the prediction of different models on the all and no overlap holdouts (**Figure 2, Supplementary Table 1**).

### Genome Wide SNP Scores

Used 1000 genomes phase III downloaded on 03/03/2016 from ftp://ftp.1000genomes.ebi.ac.uk/vol1/ftp/release/20130502/. Removed all variants that were not SNPs or that did not have rsID, leaving 78,017,615 SNPs. Calculated scores for individual SNPs via bedtools intersect using bed files used to make models, and recalculated gene expression annotation using single SNPs instead of loci. We then identified SNPs ≥ FDR 99 threshold for each model, excluding all SNPs used as positives in model building (**Supplementary Table 6**).

To calculate enrichment in the most recent blood traits GWAS^34^, we used the fGWAS^39^ platform. We downloaded the trans-ethnic MR-MEGA summary statistics for RBC, HCT, MCV, RDW, Neutro, Mono, Baso, Eosin, PLT and MPV phenotypes from http://www.mhi-humangenetics.org/en/resources/ on 08/13/21. We filtered these results to remove variants at the same genomic coordinates, keeping the most common variant and calculated nearest distance to an FDR < 0.01 variant in the corresponding TSABL model (RBC: RBC, HCT, MCV, RDW; WBC: Neutro, Mono, Baso, Eosin; platelets: PLT, MPV). We ran fGWAS using distance intervals of 25, 50 and 500 kb (ranging from most to least stringent) and found 25kb to have the best enrichment for all phenotypes (**Supplementary Figure 5**)

We used the Genomic Regions Enrichment of Annotations Tool (GREAT)^35^ to interpret these scores using version 4.0.4 found at http://bejerano.stanford.edu/great/public/html/index.php. SNPs with FDR < 0.01 as described above were submitted using the whole genome as background to assess genomic enrichment. We found that the tool did not function if more than 150,000 SNPs were submitted, so for datasets with > 150,000 SNPs (ECG, WBC), a random subset of 150,000 was submitted. To assess if enrichments changed when variants linked to known GWAS variants were removed, we removed any variants within 500 KB of a lead GWAS hit for the trait, again submitting a random 150,000 if the SNP list exceeded (ECG) this threshold. For all analysis, we recorded the GO biological processes and mouse single KO phenotype results. We note that the analyses that returned no results were all assessing < 7,000 SNPs, and expect that they failed due to having too few input regions. Full lists of variants and results can be found in **Supplementary Table 7**.

### Credible Sets

We used available summary statistics (**Supplementary Table 1**) to calculate credible sets for GWAS of interest. We limited our analysis to loci used for modelling where the lead SNP was present in the summary statistic data (this eliminated ECG, for which no summary statistics were available, and lipids, which was missing summary statistics for many lead variants). We identified variants of interest within a 1 MB window surrounding lead SNPs, limiting to variants with R^2^ ≥ 0.5 to the lead variant in European 1000 genomes phase III^44^ data using PLINK 1.90Beta4.5^45^ These variants were assessed using R package corrcov^53^ to create 95% confidence interval credible sets (**Supplementary Table 8**). To calculate percentage of credible sets our models reduce SNP number for, at each FDR threshold we found the number of credible sets where that threshold yielded at least 1, but less than the maximum number of SNPs and divided by the number of credible sets with more than 1 SNP.

### LDSC Partitioned Heritability

Downloaded LD Scores v2.2, plink files, weights, and other necessary files from https://data.broadinstitute.org/alkesgroup/LDSCORE. Added 6 annotations corresponding to 0-1 normalized SNP scores, one per trait model (autoimmune, ECG, lipids, platelets, RBC, WBC) to V2.2 annot files. Removed 11,608 SNPs on chromosome 12 containing ssIDs instead of rsIDs from both annot and plink files. Recalculated LD Scores^29^ using new annot files as per https://github.com/bulik/ldsc/wiki/LD-Score-Estimation-Tutorial. Partitioned heritability for 20 relevant GWAS (**Supplementary Table 4**) as per https://github.com/bulik/ldsc/wiki/Partitioned-Heritability. For models with some GWAS summary statistics (autoimmune, lipids, platelets, RBC, WBC), we also removed SNPs in LD R^2^ > 0.7 with lead SNPs (**Supplementary Table 12**) from both annot and plink files, then recalculated LD Scores and repartitioned heritability to test how much heritability is driven by lead SNPs (**Supplementary Table 5**).

## Data Availability

Manuscript data is available upon request

