## Supplementary Figures for "TSABL: Trait Specific Annotation Based Locus Predictor"

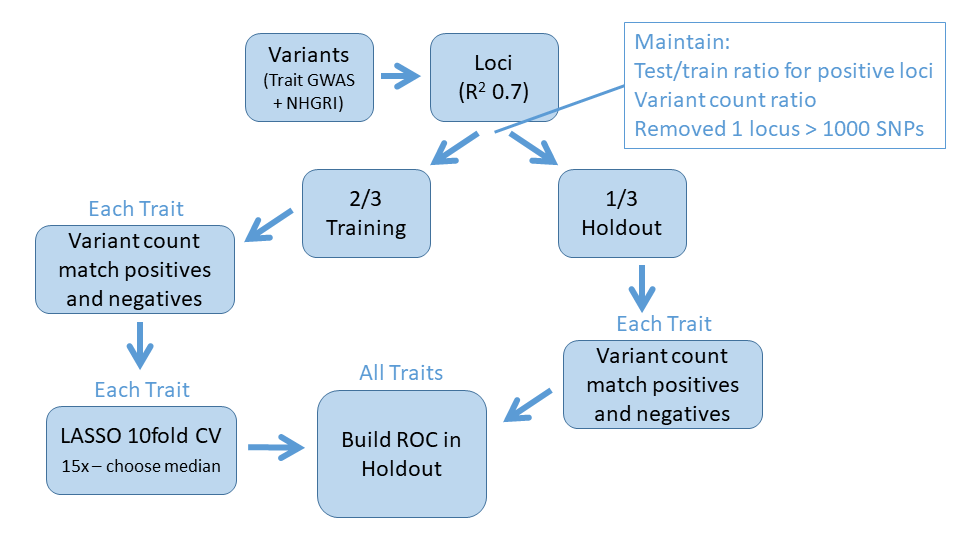

**Supplementary Figure 1: Model Building Flow Chart.**

To build models using GWAS loci, we grouped lead variants by LD, then split into training and holdout sets semi-randomly, enforcing an even distribution of positive loci and maintaining the distribution of variant counts between the sets. For modelling, we included only positive loci which had a sufficient number of negative loci with similar locus variant count, which was enforced in both training and holdout sets. After models were built, they were assessed by building ROC plots in the holdout sets.

A)	B)

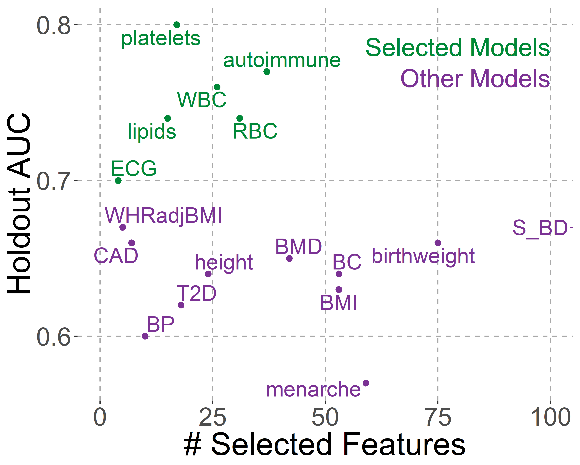

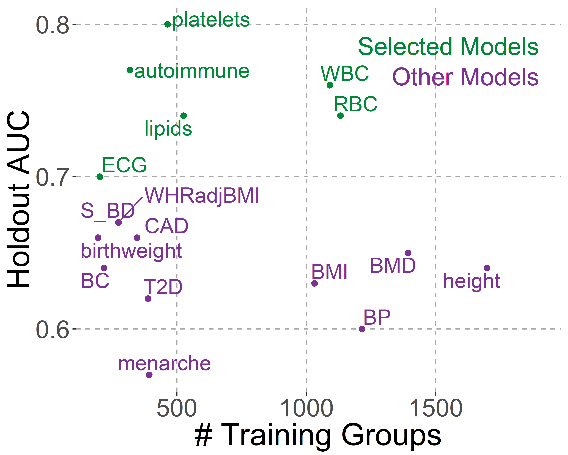

**Supplementary Figure 2: Holdout AUCs vs # Selected Features and # Groups used for Training.**

Model AUC in holdout set plotted against A) number of features selected by the model and B) number of positives groups used for training. In both panels, models which passed selection criteria (AUC ≥ 0.7, AUC change between training and holdout ≤ 0.02) are green while models which were not selected are purple.

A)

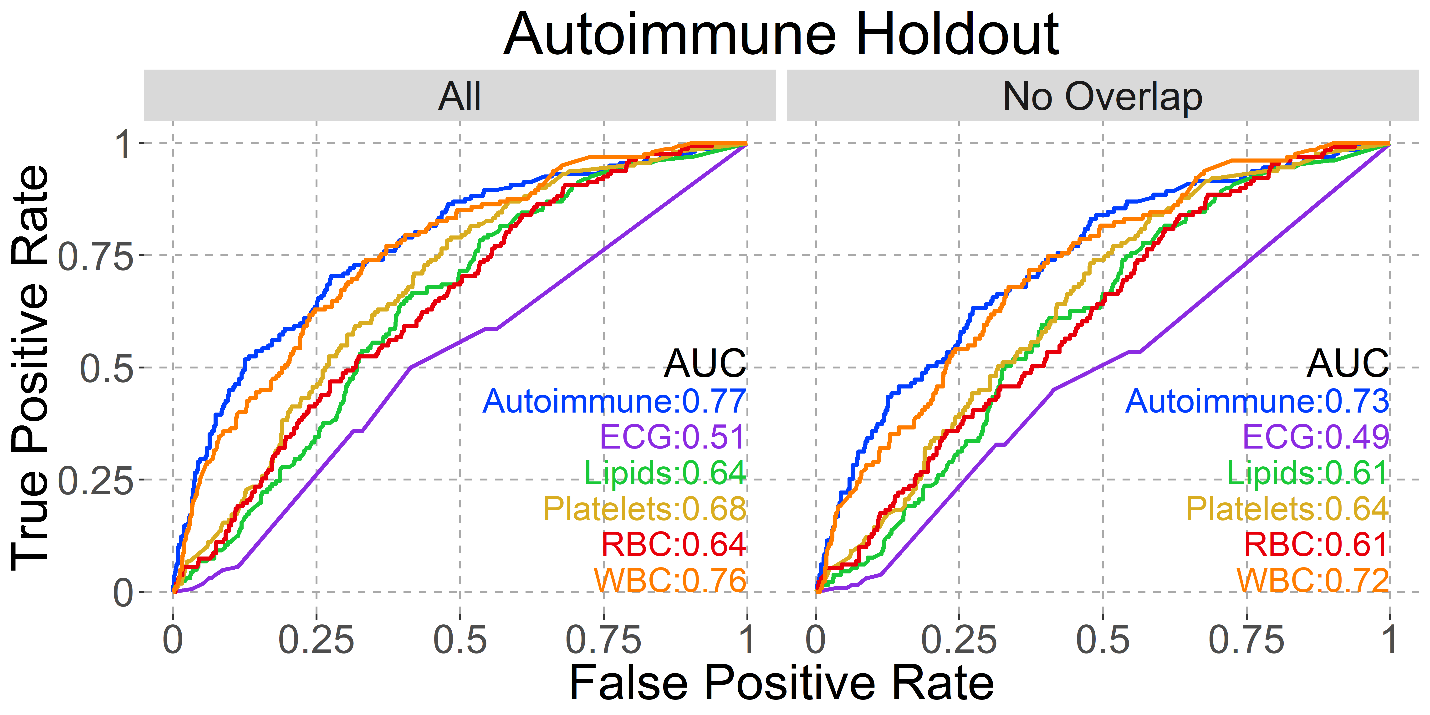

B)
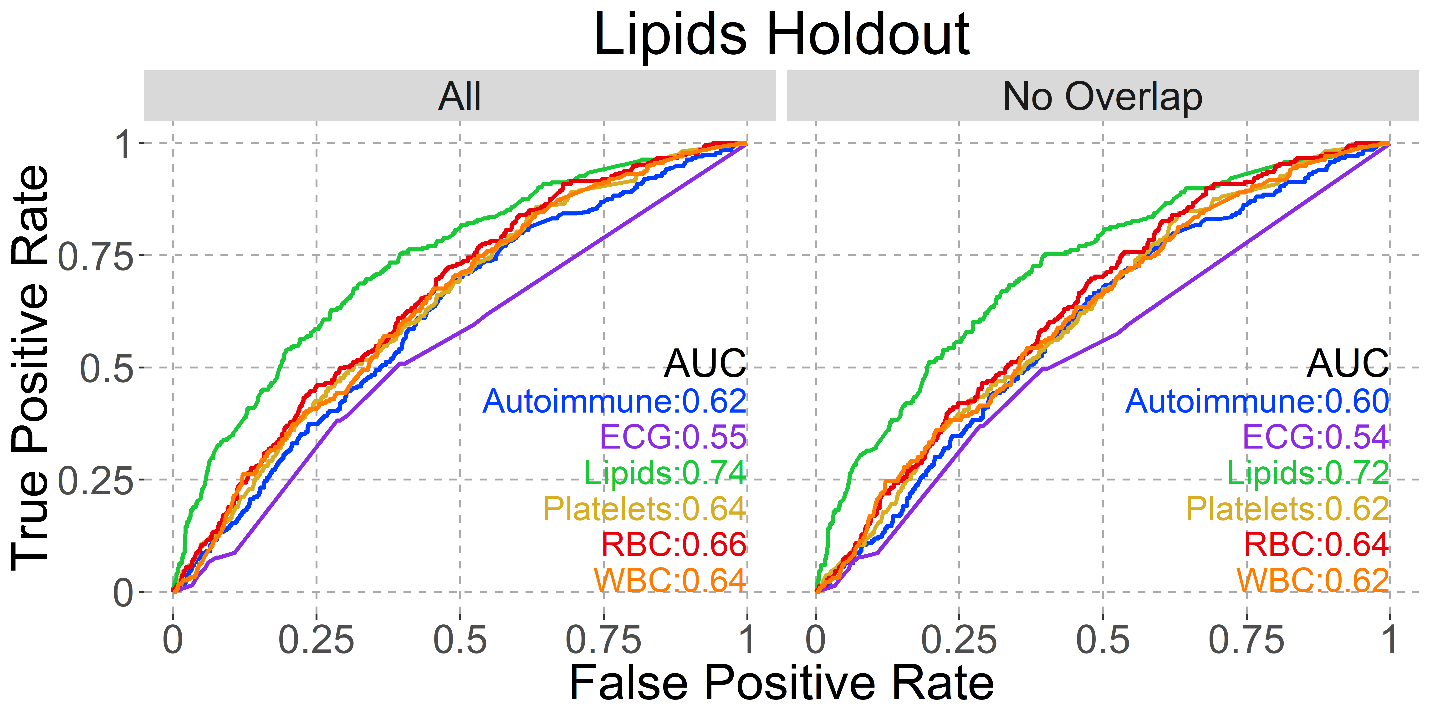

C)
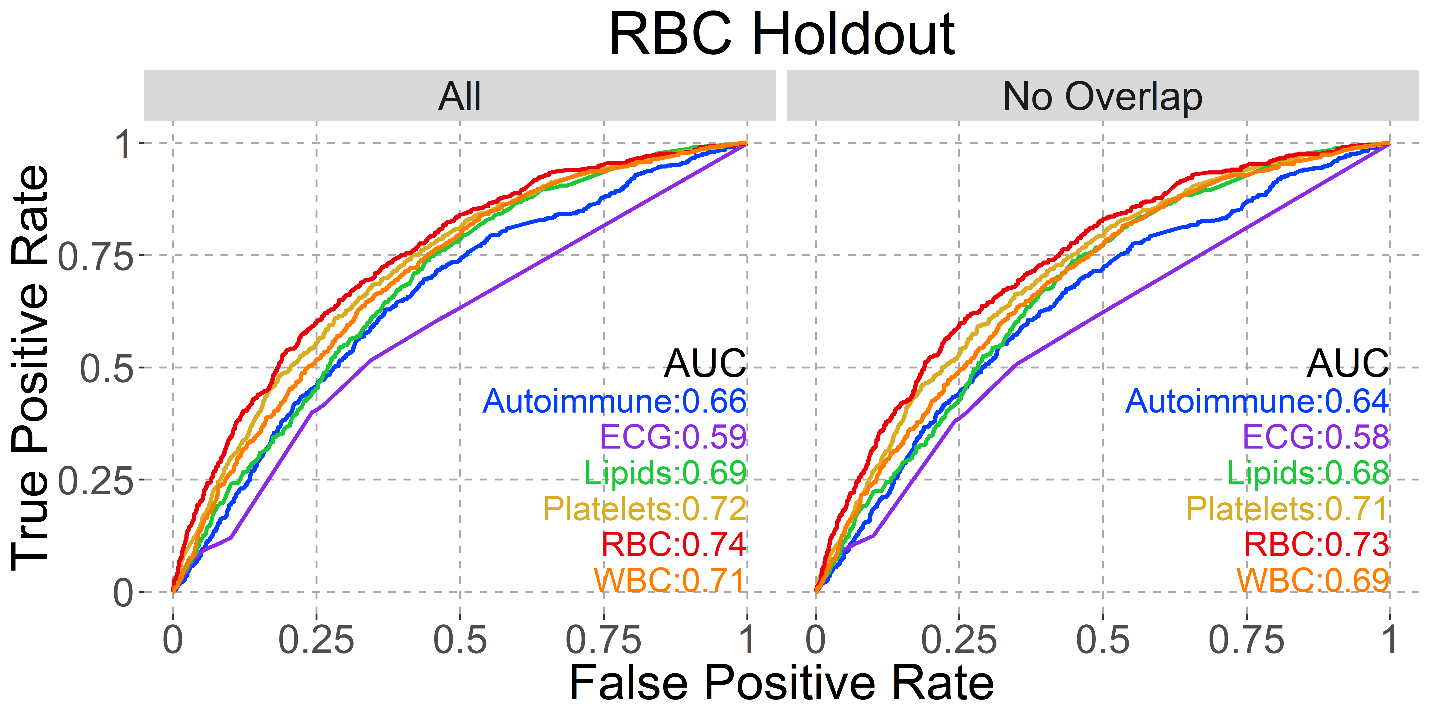

D)
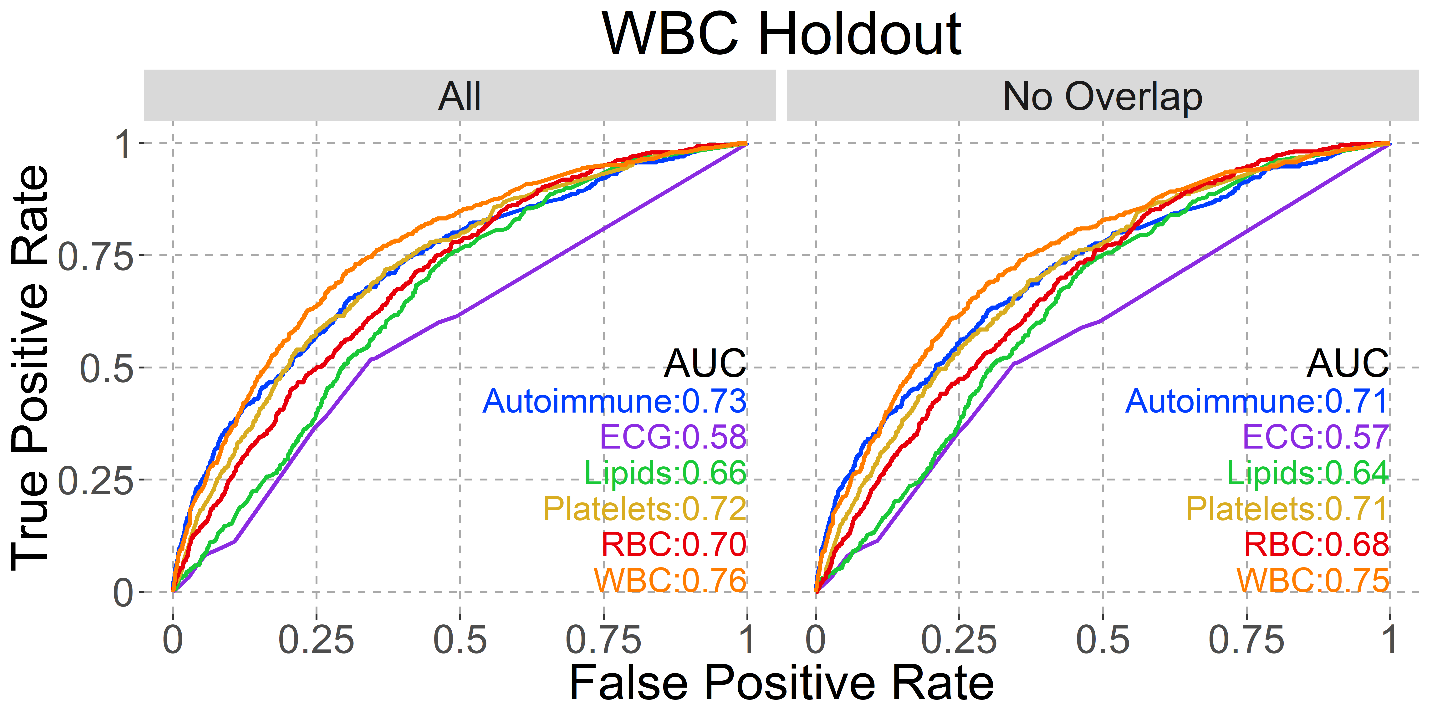

**Supplementary Figure 3: Model AUCs on same holdout sets.**

Each panel shows the named holdout set: A) Autoimmune, B) Lipids, C) Red Blood Cells and D) White Blood Cells. AUCs for all six selected models are shown on all plots, with the left (All) plot having all loci in that holdout and the right (No Overlap) having only those loci not positive in multiple models.

A)
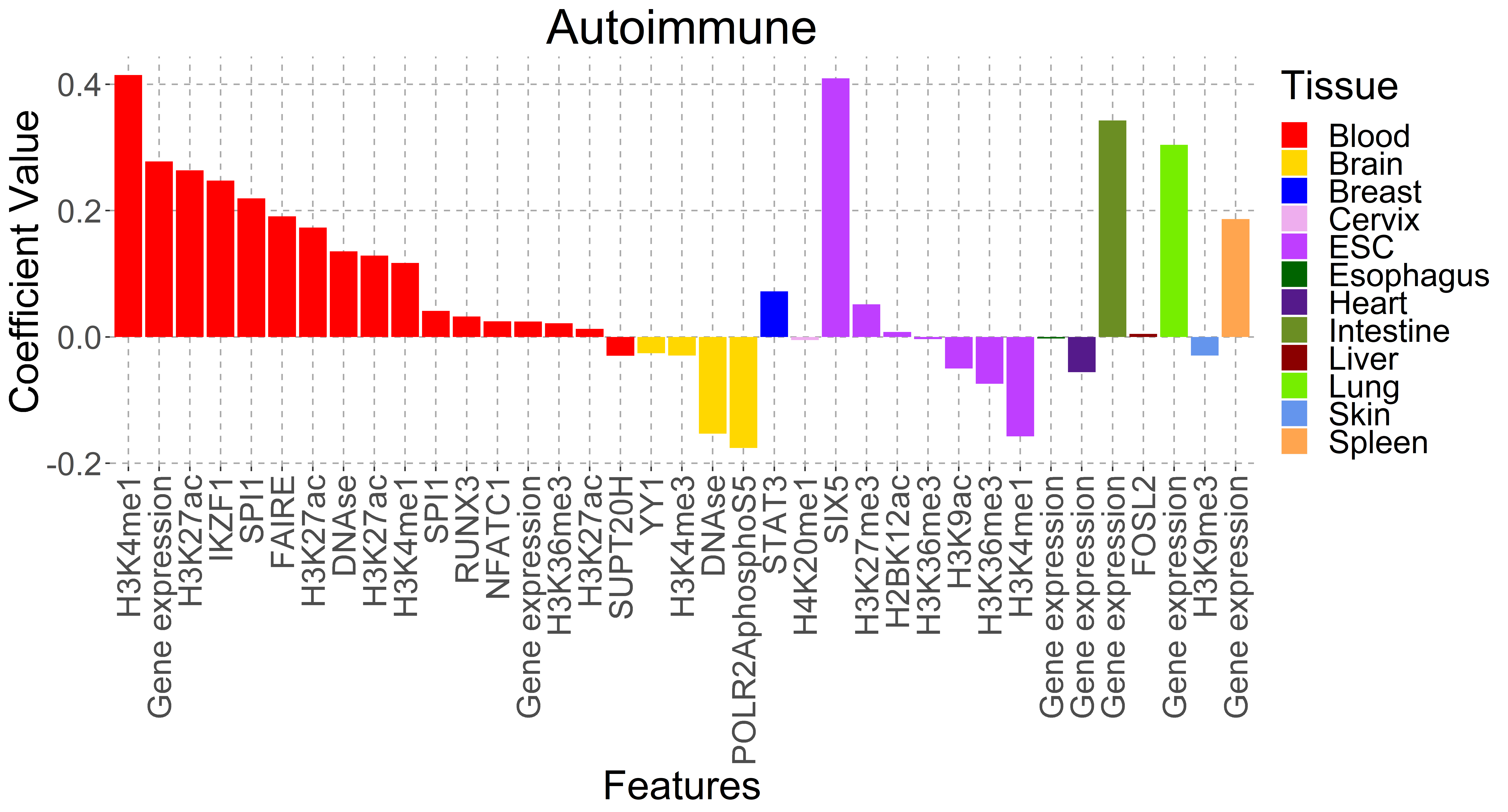
B)

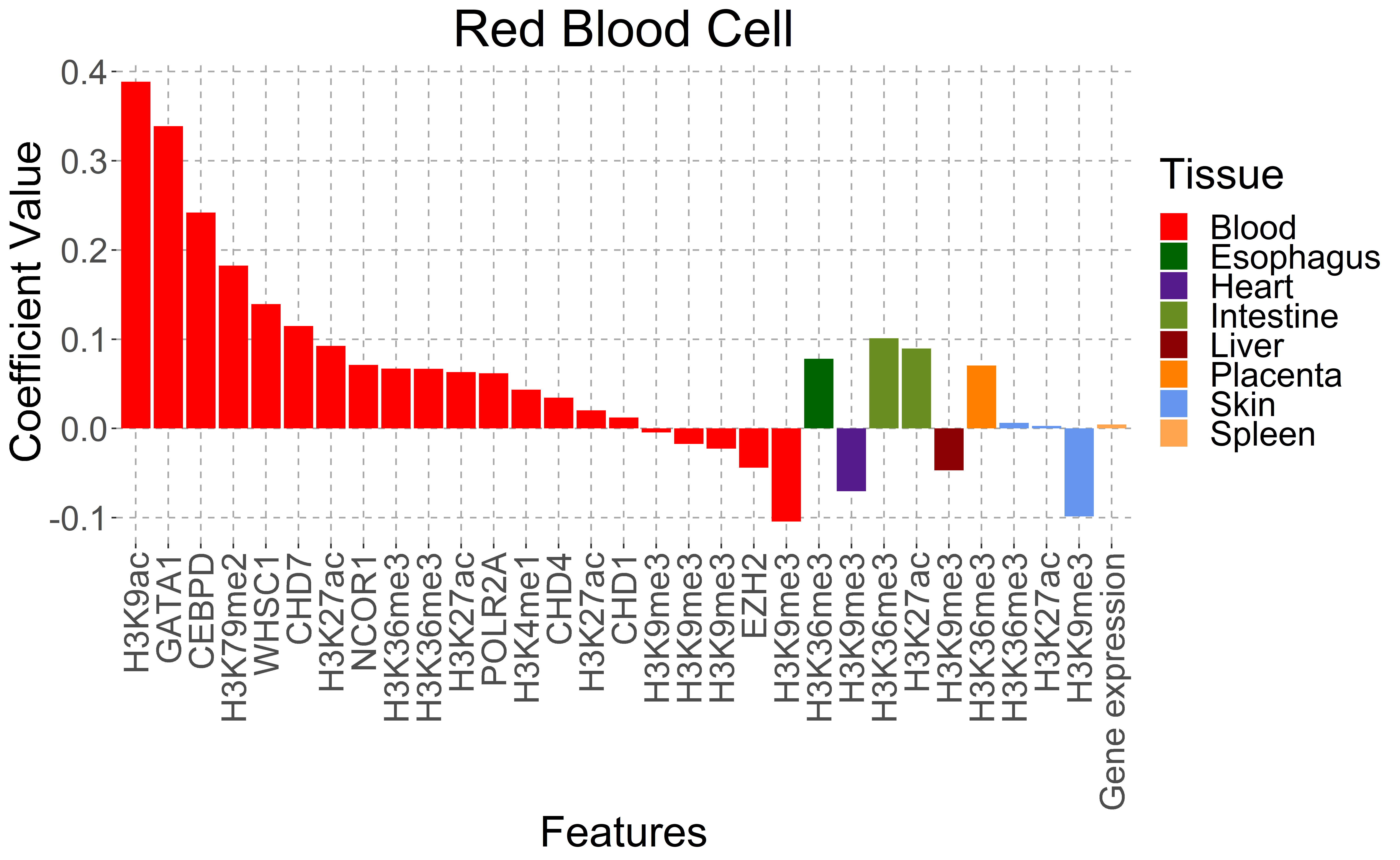

C)

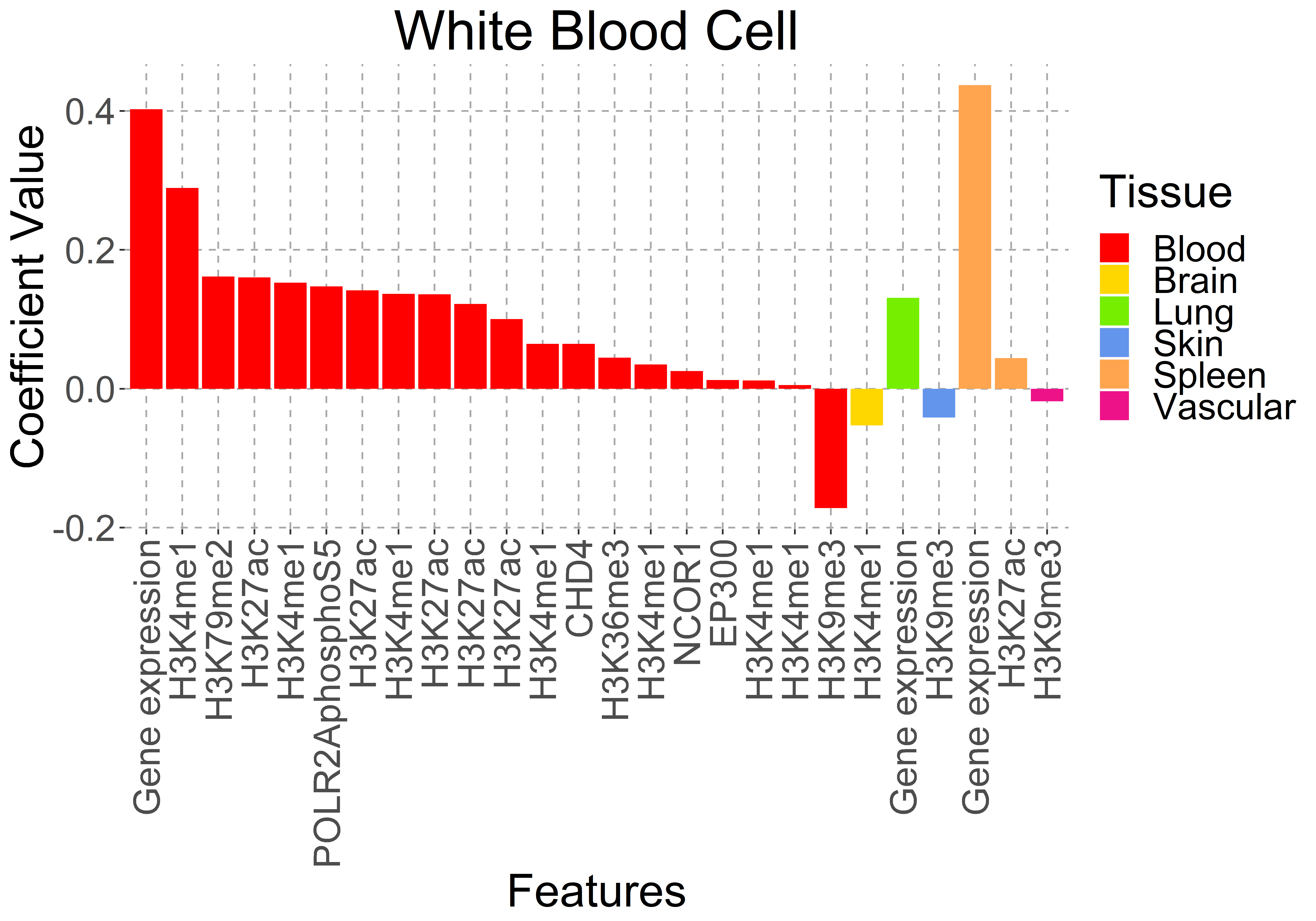

**Supplementary Figure 4: Selected Model Feature Coefficients.**

A) Autoimmune, B) Red Blood Cells, and C) White Blood Cells model coefficients, sorted by tissue type and value.

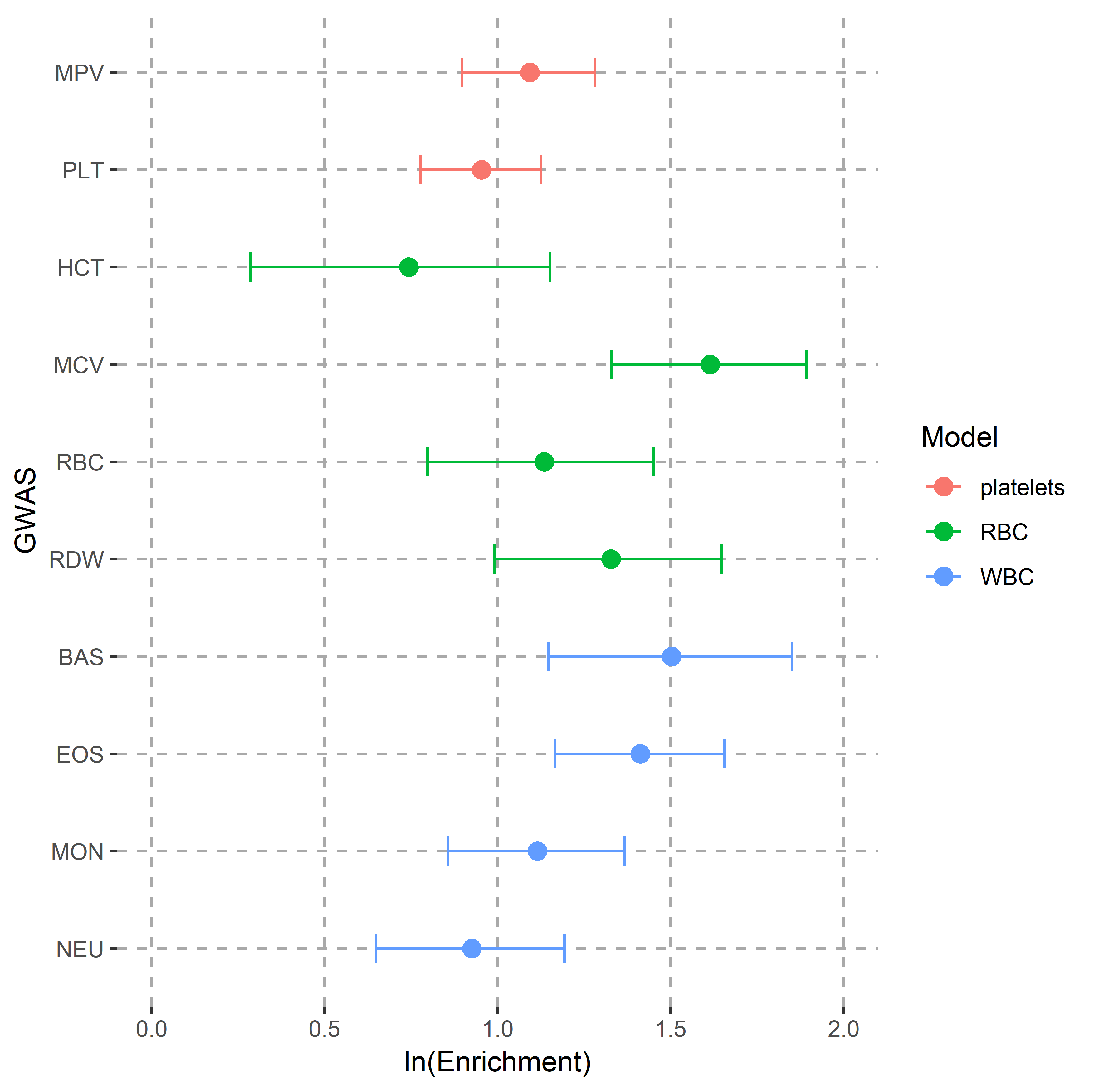

**Supplementary Figure 5: Enrichment of FDR < 0.01 SNPs within 25kb of GWAS variants.**

GWAS tested for platelets model: mean platelet volume (MPV) and platelet count (PLT); for RBC model: hematocrit (HCT), mean corpuscular volume (MCV), red blood cell count (RBC), and red cell distribution width (RDW); for WBC model: basophil count (BAS), eosinophil count (EOS), monocyte count (MON) and neutrophil count (NEU).
